# Large Language Models in Stroke Management: A Review of the Literature

**DOI:** 10.1101/2025.06.28.25330477

**Authors:** Shelly Soffer, Aya Mudrik, Orly Efros, Mahmud Omar, Girish N Nadkarni, Eyal Klang

## Abstract

Stroke care generates vast free-text records that slow chart review and hamper data reuse. Large language models (LLMs) have been trialed as a remedy in tasks ranging from imaging interpretation to outcome prediction. To assess current applications of LLMs in stroke management, we conducted a narrative review by searching PubMed and Google Scholar databases on January 30, 2025, using stroke- and LLM-related terms. This review included fifteen studies demonstrating that LLMs can: (i) extract key variables from thrombectomy reports with up to 94% accuracy, (ii) localize stroke lesions from case-report text with F1 scores of 0.74–0.85, and (iii) forecast functional outcome more accurately than legacy bedside scores in small pilot cohorts.

These results, however, rest on narrow, retrospective datasets-often from single centers or publicly available case reports that the models may have encountered during pre-training. Most evaluations use proprietary systems, limiting reproducibility and obscuring prompt design. None stratify performance by sex, language, or socioeconomic status, and few disclose safeguards against hallucination or data leakage.

We conclude that LLMs are credible research tools for text mining and hypothesis generation in stroke, but evidence for clinical deployment remains preliminary. Rigorous, multisite validation, open benchmarks, bias audits, and human-in-the-loop workflows are prerequisites before LLMs can reliably support time-critical decisions such as thrombolysis or thrombectomy triage.

## INTRODUCTION

Modern stroke care spans from symptom recognition and imaging to endovascular procedures and rehabilitation.^1^ This range produces large amounts of clinical text: notes, radiology reports, procedure logs, and discharge summaries.^2^ Large language models (LLMs) can integrate these data at scale. They may reveal clinical associations that remain hidden when documentation is scattered.^3^

LLMs may also automate time-consuming charting, giving neurologists more time for direct patient care.^4^ Properly trained models may detect high-risk stroke presentations, identify subtle imaging findings in free-text reports, or adapt patient education to diverse linguistic and cultural contexts.^5–8^ Despite these possibilities, questions remain about how LLMs will perform in varied healthcare settings, how they will handle missing or irregular data, and how clinicians can guard against errors or model “hallucinations”.^9^ These concerns are critical in stroke, where decisions often hinge on details that emerge under severe time constraints.^10^

In this review we outline current LLM applications in stroke management, as presented in **Figure 1**. We begin by clarifying basic LLM terminology, then explore their use across stroke management. We conclude with an assessment of limitations, practical considerations, and technical underpinnings.

**Figure 1.**
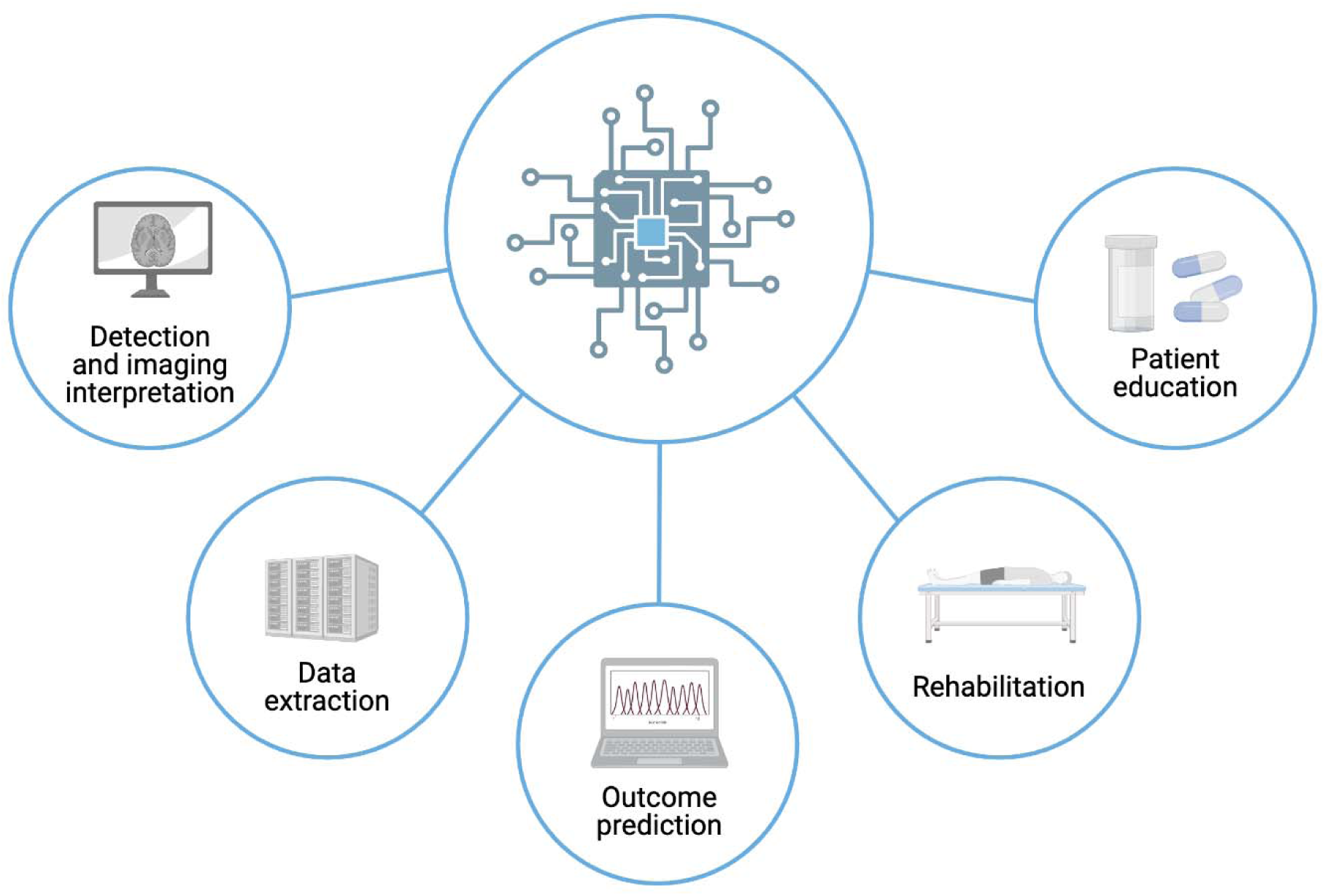
Applications of Large Language Models (LLMs) in Stroke Management.

## AI GLOSSARY OF TERMS

To establish a shared foundation for the sections that follow, we provide a concise glossary clarifying key concept such as artificial intelligence (AI), natural language processing (NLP), and large language models (LLMs). **Figure 2** presents a hierarchy diagram of AI terms.

**Figure 2.**
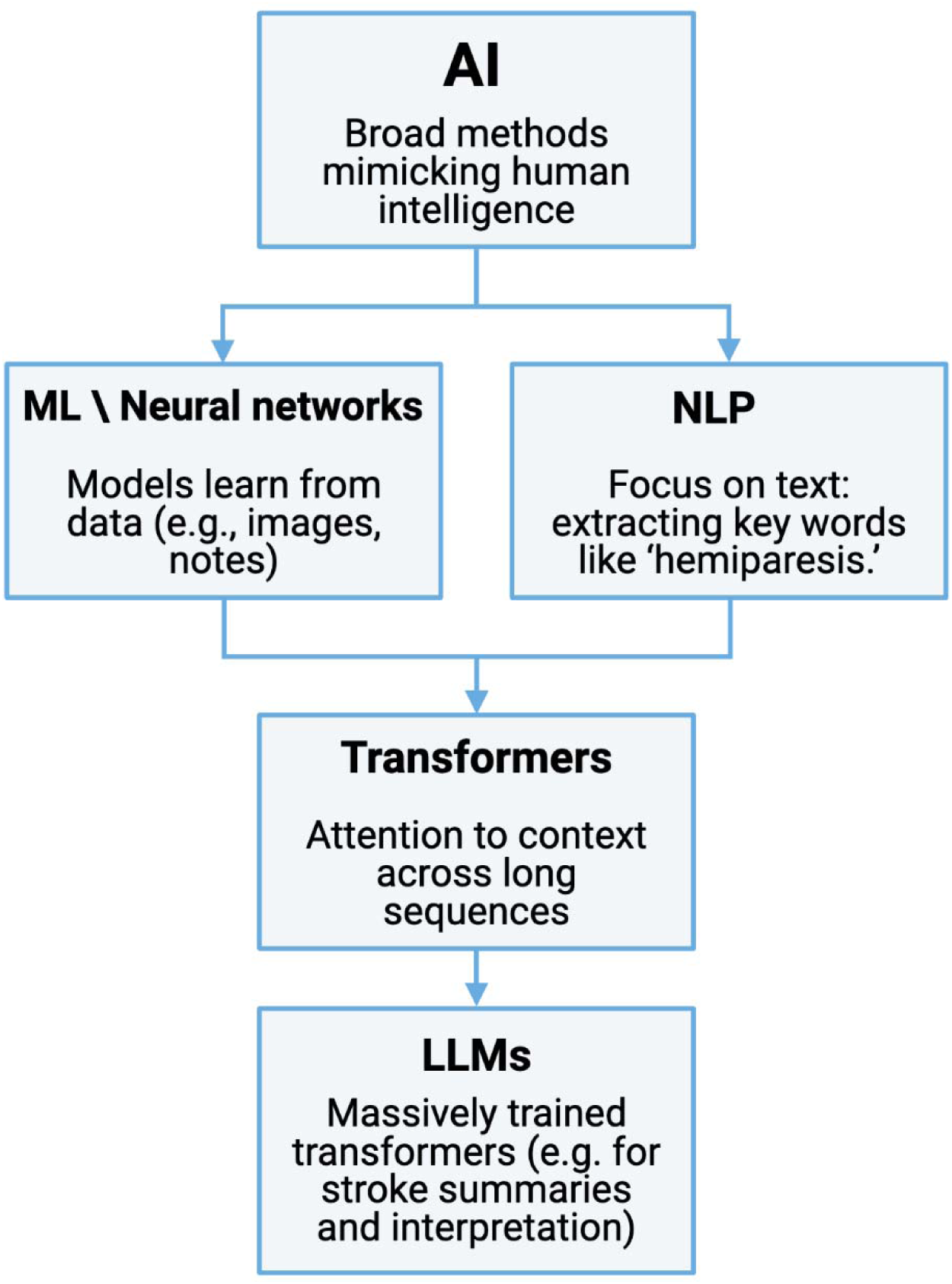
A hierarchy diagram of AI terms.

### Artificial Intelligence (AI)

AI encompasses computational methods that perform tasks traditionally requiring human judgment. ^9,11^ In stroke care, AI tools can analyze imaging findings, vital signs, and clinical text.^12,13^ This accelerates detection of critical events and may improve patient triage. AI requires careful validation to ensure reliable outputs.^14^

### Machine Learning (ML) and Neural Networks

Machine learning is a branch of AI that uses algorithms to discover patterns in data and refine them with training. ^15^ ^16^ In stroke contexts, ML models can detect subtle indications of early infarction or predict patient outcomes based on risk factors.^12,17^

Neural networks are a common ML approach, inspired by the organization of neurons in the human brain. They process data through multiple layers of connected nodes. Each “artificial neuron” is similar to one logistic regression unit, weighting inputs from the previous layer, and outputs the result to the next layer.^15,18,19^ In stroke research, such networks can classify CT or MRI findings, and pinpoint factors linked to large-vessel occlusion or hemorrhage.^20^

### Natural Language Processing (NLP)

NLP enables AI systems to interpret and generate human language. It transforms clinical notes— like admission notes or discharge summaries—into analyzable information.^21,22^ In stroke care, NLP can flag references to symptoms such as “facial droop” or “speech difficulty”.

This approach allows rapid, automated extraction of relevant patient details. Clinicians receive concise summaries of key findings, reducing manual chart review.^23^

### Transformer Architecture and Attention

Transformers are an advanced type of neural network that process sequences, such as free-text notes, by assigning weights to different words. This “attention” mechanism highlights important words while preserving context.^24^ For example, a transformer may focus on terms like “hemiparesis” or “neglect” in a clinical note.

This architecture manages large datasets efficiently. It can prioritize urgent cases by identifying high-risk descriptors in patient histories. In stroke units, where rapid decision-making matters, a transformer’s ability to distill key textual details can improve workflow.

### Large Language Models (LLMs)

LLMs are very large transformer-based models trained on extensive text corpora. They generate context-sensitive responses by predicting the next token in a sequence. In stroke care, an LLM might synthesize patient symptoms and risk factors to suggest possible diagnoses or management steps. Examples of LLMs include closed commercial chatbot models such as openAI’s GPT-4o,^25^ GPT-o1,^26^ and Anthropic’s Claude-3.5,^27^ and open access models such as Meta’s Llama-3.3,^28^ and DeepSeek R1.^29^

### Fine-Tuning

Fine-tuning adapts a pre-trained model to a targeted task using additional examples from the desired domain.^30^ In stroke research, a general LLM can be fine-tuned with registries of acute ischemic or hemorrhagic stroke cases. This process reduces errors and improves consistency.^31^

### Prompt Engineering

Prompt engineering involves designing specific queries or instructions for an LLM.^32^ Precise prompts yield more focused, accurate responses.^33^ This technique ensures that AI outputs remain clinically relevant. In practice, a well-constructed prompt can prevent the model from generating off-topic content, saving time in emergency or inpatient settings where clarity is key.^34,35^ **Figure 3** demonstrates a prompt engineering workflow for stroke diagnosis using LLMs.

**Figure 3.**
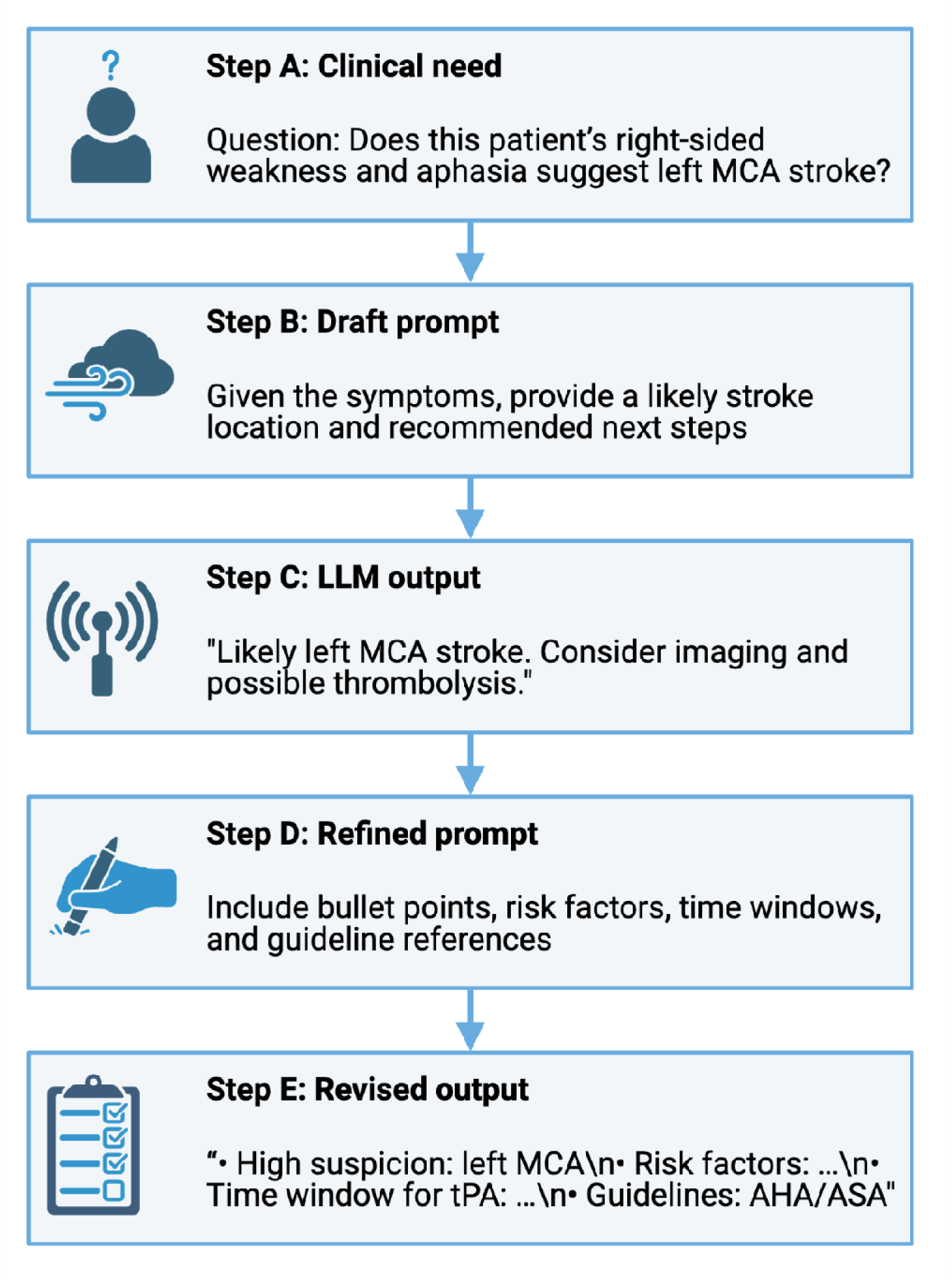
A prompt engineering workflow for stroke diagnosis using LLMs.

### Hallucinations

An LLM “hallucination” occurs when the model confidently produces content that is factually incorrect or unfounded.^36^ Several factors can cause hallucinations, including incomplete training data or ambiguous prompts. Continuous monitoring and human oversight are important. Regular testing of the model’s outputs using real-world scenarios helps maintain reliability. ^37^

## METHODS

This review followed general principles for narrative reviews as outlined by Green et al.^38^ and Ferrari^39^. We conducted a narrative review to explore the applications of LLMs in the context of stroke. Our literature search was performed on January 30, 2025 using key terms related to both stroke and LLMs across PubMed and Google Scholar. The full list of search terms is provided in the **Supplement Materials**. In addition, we screened the reference lists of all included articles to identify additional studies.

We included studies that addressed the use of LLMs in stroke care, diagnosis, intervention, or research settings, and excluded those that did not substantively engage with both domains. Article selection was based on title and abstract screening, followed by full-text review when appropriate.

From the included studies, we extracted information on study design, population characteristics, and reported use cases or outcomes. We organized the findings thematically, emphasizing clinical applications, model functions, and methodological approaches and limitations.

Consistent with narrative review methodology, we did not perform formal statistical analyses or risk-of-bias assessments. Instead, we aimed to summarize current developments, map emerging trends, and suggest avenues for future research and clinical practice.

## CLINICAL APPLICATIONS

We included 15 studies in this narrative review. Their characteristics, key findings, and metrics are summarized in **Tables 1–5**. Limitations and future directions are detailed in **Supplementary Tables 1–5**.

**Table 1:**
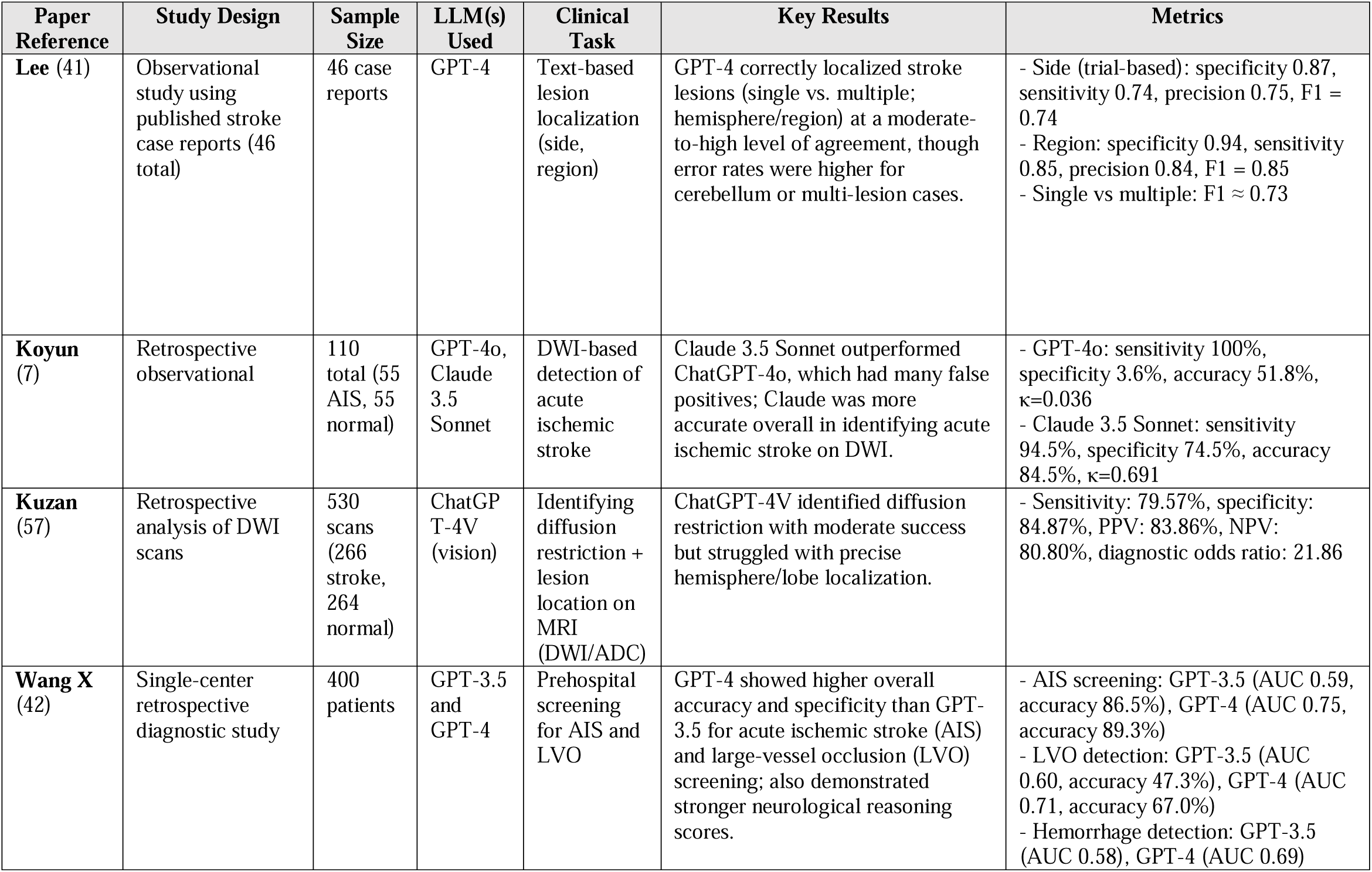
Examples of LLM applications in stroke detection and imaging.

### Stroke Detection and Imaging Interpretation

Recent investigations highlight how LLMs can assist in stroke detection and imaging interpretation, with direct implications for timely care.

For example, Lee et al. evaluated openAI’s GPT-4^40^ for localizing acute stroke lesions using raw text from 46 published case reports (BMC Neurology case reports).^41^ They asked GPT-4 to determine whether each case involved a single vs. multiple lesion, which brain region (cerebrum, cerebellum, brainstem, or spinal cord), and which side (left, right, or both). Each case was tested three times. Then, GPT-4’s answers were compared with the actual imaging results. Across these 138 trials, GPT-4 achieved F1-scores of about 0.74–0.85 for region/side detection, performing well for cerebral and spinal lesions but struggling more with cerebellar cases. Some errors stemmed from incomplete or ambiguous case descriptions (“extrinsic”), while others reflected GPT-4’s own logical slips (“intrinsic”). One notable limitation is that the cases were published, potentially giving GPT-4 prior exposure to them during its training. Also, case reports often omit routine details and may feature rare presentations, so results may not generalize to everyday clinical notes.

LLMs have also demonstrated promise in assisting emergency medical services (EMS) by enhancing prehospital screening tools. The recent retrospective study of 400 emergency department patients by Wang et al. evaluated GPT-4 and GPT-3.5 for the detection of acute ischemic stroke (AIS) and large vessel occlusion (LVO).^42^ GPT-4 achieved a higher area under the receiver operating characteristic curve (AUC) for AIS (0.75 vs 0.59) and LVO detection (0.71 vs 0.60), alongside superior sensitivity and specificity for both conditions. Moreover, GPT-4 demonstrated stronger factual correctness (Likert score of 4.24 vs 3.62) and a lower rate of errors (6.8% vs 24.8%) compared to GPT-3.5.

Koyun and Taskent retrospectively examined 110 Diffusion-Weighted (DW) MRI cases—55 with AIS and 55 healthy controls—to compare the diagnostic performance of two AI models, openAI’s GPT-4o^25^ and Anthropic’s Claude 3.5 Sonnet^27^, in detecting AIS.^7^ While GPT-4o achieved an accuracy 51.8%, Claude 3.5 Sonnet showed a substantially higher accuracy 84.5%. In hemisphere-level localization, Claude 3.5 Sonnet was correct in 67.3% of AIS cases, whereas GPT-4o succeeded in 32.7%. For specific region/lobe localization, Claude 3.5 Sonnet’s accuracy reached 30.9%, while GPT-4o’s was 7.3%. A second evaluation two weeks later showed moderate intra-model agreement for both, but Claude 3.5 Sonnet consistently outperformed GPT-4o. The authors conclude that Claude 3.5 Sonnet demonstrates higher reliability for AIS detection and localization than GPT-4o, though both models exhibit limitations that underscore the need for further refinement before routine clinical use.

**Table 1** and **Supplementary Table 1** present examples of LLM applications in stroke detection and imaging interpretation.

### Data extraction

One of the big advantages of NLP and specifically LLMs is extracting structured data from free-text. Critical data points are frequently buried in notes or reports, making them difficult to search and analyze. By automatically converting free-text information into structured formats—such as standardized fields for demographics, imaging results, or procedural timestamps—clinicians can quickly access key insights, identify patterns, and integrate these data into stroke registries or prediction models.^43^

Lehnen et al. conducted a retrospective analysis of 130 free-text neuroradiology reports (100 internal and 30 external) from mechanical thrombectomy procedures, comparing GPT-4 and GPT-3.5 on their ability to automatically extract standardized procedural details. Their results showed that GPT-4 correctly retrieved 94% of data points (e.g., occlusion site, times, and materials used), substantially outperforming GPT-3.5 (64%) in both the main and external datasets.^44^ Notably, prompt refinements further improved GPT-4’s performance on challenging fields like “last thrombectomy maneuver time,” underscoring the importance of prompt design.

Similarly, Meddeb et al. evaluated three locally deployed open-source LLMs—Mistral’s Mixtral,^45^ Alibaba’s Qwen,^46^ and BioMistral fine-tuned model^47^—on more than 1000 mechanical thrombectomy reports, testing their ability to extract 15 clinical data fields (e.g., National Institutes of Health Stroke Scale [NIHSS] scores, occluded vessels, medication details).^48^ Mixtral demonstrated particularly high precision (up to 0.99 for certain time metrics), while Qwen and BioMistral showed variable but still useful performance across different data fields. In addition, a human-in-the-loop (HITL) approach reduced the time required for final data labeling by over 65%. This underscores how partial automation can significantly expedite documentation without sacrificing accuracy. The results highlight the scalability of open-source LLMs for large-volume data extraction Goh et al. examined the feasibility of using a locally deployed LLM (Meta’s Llama 3)^28^ to automate stroke audit data extraction from free-text discharge summaries. By comparing the model’s outputs to a human-maintained statewide stroke registry, they found a 93.8% accuracy rate across multiple data fields (e.g., wake-up stroke status, past medical history, pre-stroke medications), highlighting the potential of LLMs to streamline and improve consistency in audit processes.^6^

**Table 2** and **Supplementary Table 2** present examples of LLM applications in stroke data extraction.

**Table 2:**
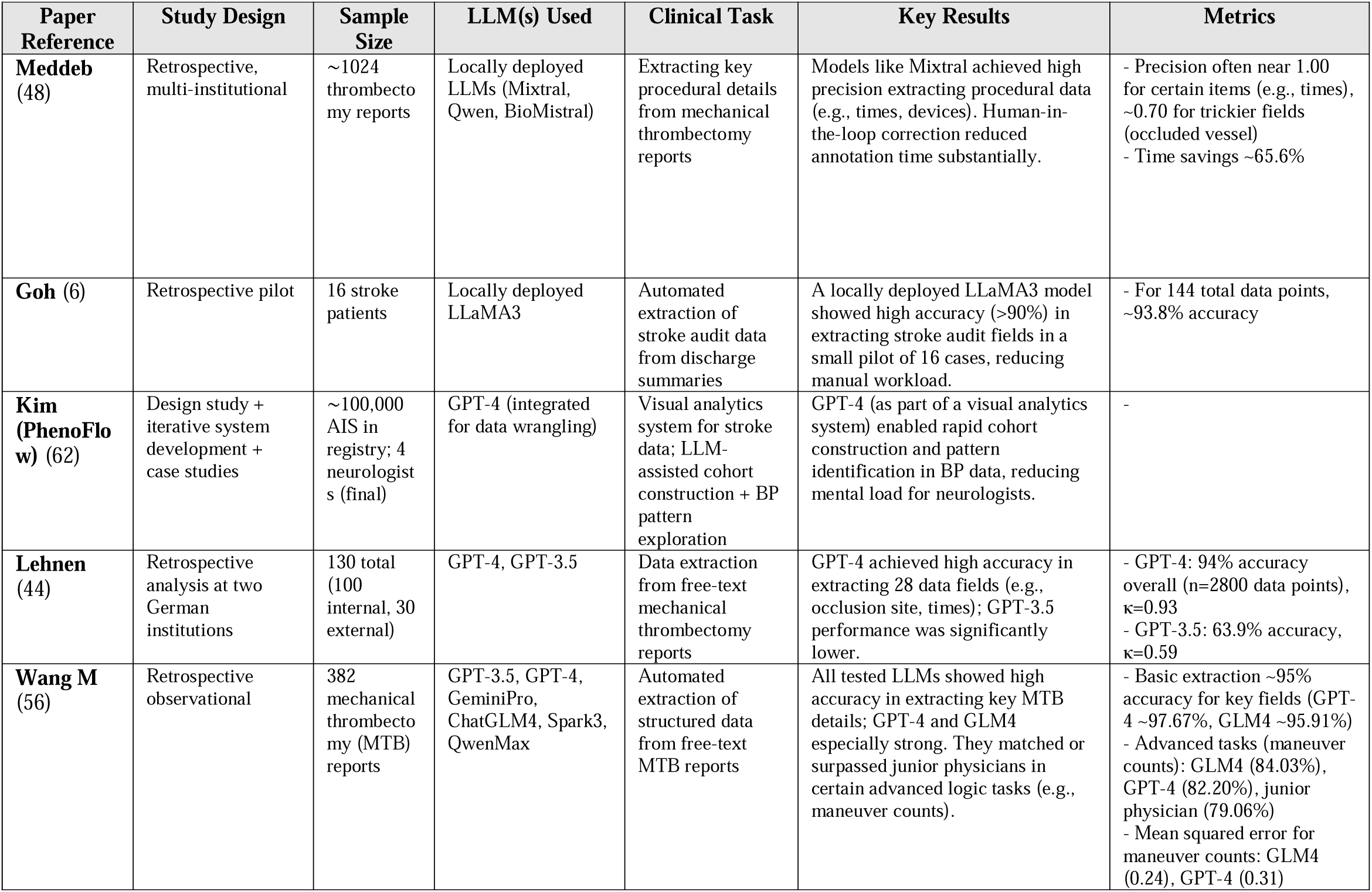
Examples of LLM applications in stroke data extraction.

| Paper Reference | Study Design | Sample Size | LLM(s) Used | Clinical Task | Key Results | Metrics |
| --- | --- | --- | --- | --- | --- | --- |
| **Meddeb (48)** | Retrospective, multi-institutional | ~1024 thrombectomy reports | Locally deployed LLMs (Mixtral, Qwen, BioMistral) | Extracting key procedural details from mechanical thrombectomy reports | Models like Mixtral achieved high precision extracting procedural data (e.g., times, devices). Human-in-the-loop correction reduced annotation time substantially. | - Precision often near 1.00 for certain items (e.g., times), ~0.70 for trickier fields (occluded vessel)<br>- Time savings ~65.6% |
| **Goh (6)** | Retrospective pilot | 16 stroke patients | Locally deployed LLaMA3 | Automated extraction of stroke audit data from discharge summaries | A locally deployed LLaMA3 model showed high accuracy (>90%) in extracting stroke audit fields in a small pilot of 16 cases, reducing manual workload. | - For 144 total data points, ~93.8% accuracy |
| **Kim (PhenoFlow) (62)** | Design study + iterative system development + case studies | ~100,000 AIS in registry; 4 neurologists (final) | GPT-4 (integrated for data wrangling) | Visual analytics system for stroke data; LLM-assisted cohort construction + BP pattern exploration | GPT-4 (as part of a visual analytics system) enabled rapid cohort construction and pattern identification in BP data, reducing mental load for neurologists. | - |
| **Lehnen (44)** | Retrospective analysis at two German institutions | 130 total (100 internal, 30 external) | GPT-4, GPT-3.5 | Data extraction from free-text mechanical thrombectomy reports | GPT-4 achieved high accuracy in extracting 28 data fields (e.g., occlusion site, times); GPT-3.5 performance was significantly lower. | - GPT-4: 94% accuracy overall (n=2800 data points), $\kappa=0.93$<br>- GPT-3.5: 63.9% accuracy, $\kappa=0.59$ |
| **Wang M (56)** | Retrospective observational | 382 mechanical thrombectomy (MTB) reports | GPT-3.5, GPT-4, GeminiPro, ChatGLM4, Spark3, QwenMax | Automated extraction of structured data from free-text MTB reports | All tested LLMs showed high accuracy in extracting key MTB details; GPT-4 and GLM4 especially strong. They matched or surpassed junior physicians in certain advanced logic tasks (e.g., maneuver counts). | - Basic extraction ~95% accuracy for key fields (GPT-4 ~97.67%, GLM4 ~95.91%)<br>- Advanced tasks (maneuver counts): GLM4 (84.03%), GPT-4 (82.20%), junior physician (79.06%)<br>- Mean squared error for maneuver counts: GLM4 (0.24), GPT-4 (0.31) |

### Outcome Prediction

Predicting stroke outcomes is an evolving challenge, as conventional scoring systems like the MT-DRAGON score often fail to incorporate real-time, dynamic patient data.^49^ AI has emerged as a powerful alternative by integrating multimodal inputs, including clinical parameters, imaging findings, and time-dependent variables.

In a pilot study, Pedro et al. assessed ChatGPT’s performance in predicting post-thrombectomy functional outcomes, finding that it outperformed the MT-DRAGON score in forecasting modified Rankin Scale (mRS) scores at three months. Notably, AI predictions were more accurate for patients with shorter onset-to-door delays and better reperfusion status, suggesting that LLMs could refine post-stroke prognosis stratification.^50^

Phillips et al. introduce “HELMET,” a hybrid machine learning framework that combines a fine-tuned LLMs with structured electronic health record (EHR) variables to predict malignant cerebral edema in large middle cerebral artery (MCA) stroke.^51^ They trained and validated two models (HELMET-8 and HELMET-24) on a retrospective cohort of 623 patients from two Massachusetts hospitals (yielding over 15,000 patient-hour observations) and then externally tested on 60 patients at a separate safety-net hospital (over 3,700 observations). By continuously evaluating the degree of midline shift (MLS)—a critical indicator of severe edema—across 8-hour and 24-hour horizons, HELMET outperforms simpler regression-based scores and generalizes effectively to an external validation cohort. The authors aim to provide a real-time, dynamic risk stratification tool that can help clinicians monitor and intervene on malignant edema as it evolves, rather than relying on static early predictions alone.

**Table 3** and **Supplementary Table 3** present examples of LLM applications in stroke outcome prediction.

**Table 3:** Examples of LLM applications in stroke outcome prediction.

| Paper Reference | Study Design | Sample Size | LLM(s) Used | Clinical Task | Key Results | Metrics |
| --- | --- | --- | --- | --- | --- | --- |
| **Phillips (51)** | Retrospective derivation + prospective validation | 623 (derivation), 60 (validation) | Fine-tuned Transformer-based LLM (“Clinical-Longformer”) | Dynamic prediction of malignant edema (midline shift) in large MCA stroke | A hybrid model integrating LLM-derived features outperformed static risk scores in forecasting future midline shift, particularly for short-term (8–24 hr) prognostication. | - Derivation: AUROC ~96%<br>- External validation: HELMET-24 up to ~70% AUROC, HELMET-8 up to ~92.5% AUROC (significantly higher than comparator) |
| **Pedro T (50)** | Retrospective pilot | 163 AIS patients (anterior circulation) | ChatGPT (version 3.5) | Predicting 3-month modified Rankin Scale (mRS) after mechanical thrombectomy | ChatGPT-3.5 demonstrated fair-to-good agreement in predicting 3-month mRS, performing better than a traditional risk score (MT-DRAGON) for dichotomized functional outcomes. | - Exact match: ~49.1%, $\kappa=0.354$ (fair)<br>- Dichotomized (0–2 vs. 3–5): $\kappa=0.727$ (good)<br>- MT-DRAGON comparison: $\kappa=0.149$ (poor) |

### Rehabilitation and Long-Term Management

Stroke rehabilitation is a highly individualized process, yet AI-driven models may optimize therapy by predicting recovery potential and tailoring interventions. Zhang et al. explored ChatGPT-4’s ability to generate rehabilitation prescriptions and classify stroke patients according to ICF (International Classification of Functioning, Disability, and Health) codes, finding that it provided comprehensive, rational therapy plans.^52^ While minor classification errors were noted, AI’s ability to generate structured rehabilitation strategies in seconds offers an opportunity to personalize post-stroke care.

LLMs may also help with cognitive and language recovery. Cong et al. investigated AI’s application in aphasia assessment, demonstrating that pre-trained models could detect language deficits in stroke patients and even subtype aphasia based on linguistic patterns. Such models could augment traditional speech therapy, enabling automated conversational training tailored to individual deficits.^53^

**Table 4** and **Supplementary Table 4** present examples of LLM applications in rehabilitation and long-term management.

**Table 4:**
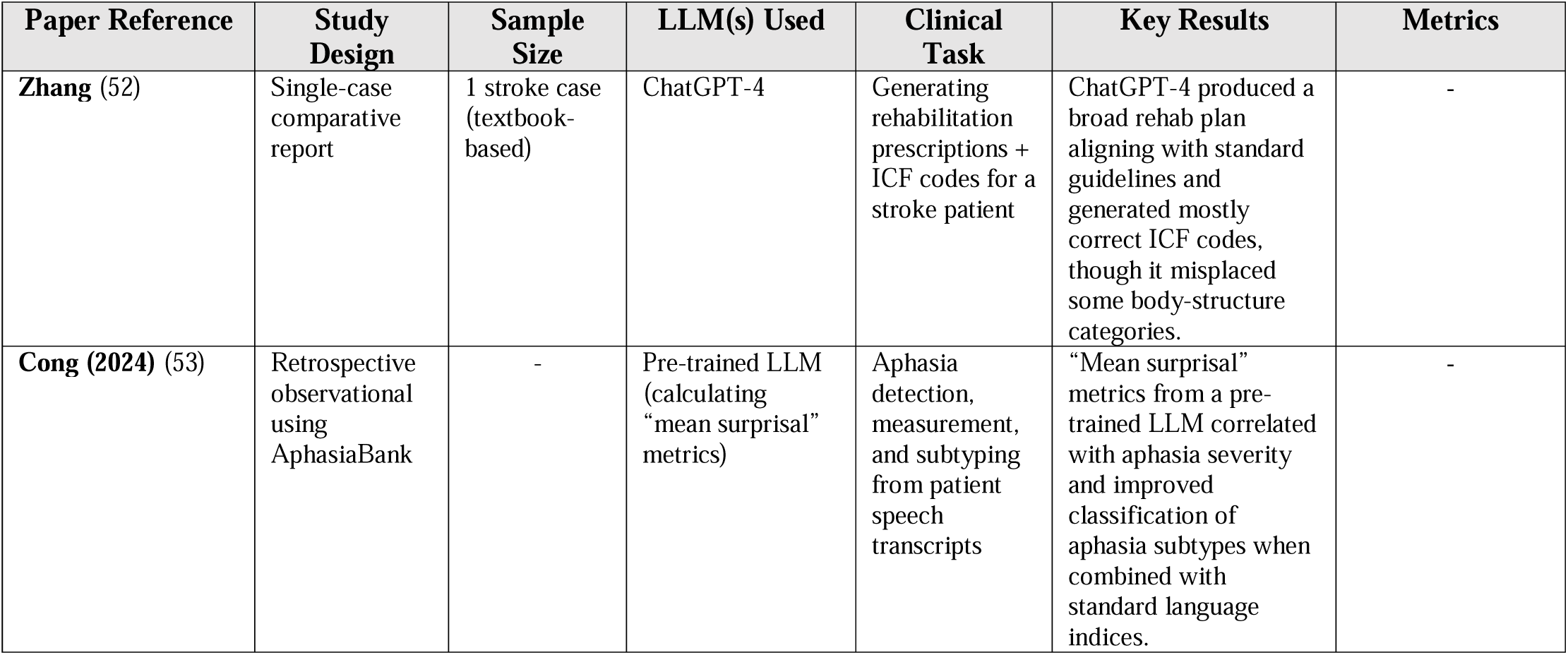
Examples of LLM applications in stroke rehabilitation.

| Paper Reference | Study Design | Sample Size | LLM(s) Used | Clinical Task | Key Results | Metrics |
| --- | --- | --- | --- | --- | --- | --- |
| Zhang (52) | Single-case comparative report | 1 stroke case (textbook-based) | ChatGPT-4 | Generating rehabilitation prescriptions + ICF codes for a stroke patient | ChatGPT-4 produced a broad rehab plan aligning with standard guidelines and generated mostly correct ICF codes, though it misplaced some body-structure categories. | - |
| Cong (2024) (53) | Retrospective observational using AphasiaBank | - | Pre-trained LLM (calculating “mean surprisal” metrics) | Aphasia detection, measurement, and subtyping from patient speech transcripts | “Mean surprisal” metrics from a pre-trained LLM correlated with aphasia severity and improved classification of aphasia subtypes when combined with standard language indices. | - |

### Patient Education

LLMs demonstrate potential in stroke-related patient education by explaining ischemic stroke pathophysiology, diagnostic work-up, and secondary prevention strategies in plain language.^54,55^ In simulated patient interactions, these models promptly advised calling emergency services, and guided users through what to expect upon arrival at the emergency department, illustrating how real-time dialogue may enhance symptom recognition and prompt decision-making.^54^ However, LLMs exhibit notable limitations, such as omitting major risk factors and therapeutic time windows,^55^ and pose practical challenges for patients with dysarthria, dysphasia, aphasia, or alexia— barriers that must be addressed before clinical integration.^54^

**Table 5** and **Supplementary Table 5** present examples of LLM applications in patient education.

**Table 5:** Examples of LLM applications in stroke patient education.

| Paper Reference | Study Design | Sample Size | LLM(s) Used | Clinical Task | Key Results | Metrics |
| --- | --- | --- | --- | --- | --- | --- |
| **Lam (54)** | Letter to editor (single-case example) | 1 conversation | ChatGPT | Immediate guidance for suspected stroke symptoms (patient-facing chatbot scenario) | ChatGPT provided advice aligned with stroke warning signs (FAST) in a single example, recommending urgent medical attention; highlights potential for immediate triage assistance but requires caution. | - |
| **Vora (55)** | Letter to the editor / commentary | — | ChatGPT | General public awareness of ischemic stroke | ChatGPT gave readable, general info on ischemic stroke for laypeople but missed some key risk factors (e.g., hypertension) and omitted treatment time windows in its explanation. | - |

## CHALLENGES AND LIMITATIONS

The following cross-cutting weaknesses emerge across the 15 studies:

### Evidence quality

Most performance claims rest on retrospective, single-center cohorts or curated case reports — settings that do not reflect the complexity and variability of real-world stroke care. These studies often lack diversity in stroke subtypes, presentation severity, and comorbidities, limiting their applicability across diverse clinical environments, particularly in EDs or telestroke settings.

### Reproducibility constraints

Nine of the fifteen studies rely solely on proprietary APIs.^7,41,42,44,50,54–57^ Neither code nor weights are publicly available, and full prompts are not always shared. This opacity prevents independent replication and stress-testing.

### Heterogeneous, incomparable benchmarks

Studies report F1 score, AUC, accuracy, or mean absolute error on different tasks such as lesion localization, outcome prediction, or registry extraction. Without shared datasets or standard endpoints, cross-study comparisons are tenuous and risk misleading readers.

### Regulatory, legal, and privacy hurdles

Most papers mention none of HIPAA, GDPR, FDA Software-as-a-Medical-Device guidance, or the EU AI Act. While LLMs may support rapid documentation or prediction, their integration into stroke workflows must comply with these legal frameworks.^58^ Deployment pathways, cybersecurity standards, and liability frameworks remain uncharted.

### Equity and bias

The studies generally did not stratify performance by sex, race, primary language, or insurance status —factors known to influence stroke outcomes.^59^ LLMs trained on historical records may perpetuate existing treatment gaps unless bias audits and mitigation steps are built in.^60,61^

### Ethical and safety concerns

Hallucinations, silent omissions, and over-confident outputs threaten patient safety.^6,42,50^ Few studies report human-in-the-loop guardrails, real-time monitoring, or fallback protocols.^48,51,62^

## DISCUSSION

In stroke care, “time is brain”. This core principle has driven developments such as in-hospital CT scanners and tele-stroke networks, all aimed at accelerating diagnosis and treatment.^63^ We may now face a new technological inflection point: integrating LLMs into everyday practice to aid stroke detection, documentation, outcome prediction, and recovery planning.

LLMs can review unstructured text from diverse sources, including real-time imaging logs and historical case reports, and may address persistent challenges in stroke care.^43^ Early GPT-4 research suggests potential in automating tasks such as extracting procedural details from thrombectomy reports or synthesizing registry data for predictive analytics. ^48^ ^51^ This could free stroke neurologists to focus on clinical judgment, patient communication, and team coordination. Preliminary evidence also hints that LLMs might forecast risks of malignant edema or long-term disability, enabling real-time predictions that adapt as a patient’s condition evolves.^51^

Rigorous validation remains critical. “Hallucinations”—where a model offers a convincing but incorrect statement—underscore the danger of relying on AI without safeguards. ^36^ ^37^ In a time-sensitive stroke activation, an erroneous conclusion could carry serious consequences. Human-in-the-loop systems that combine AI outputs with clinician expertise may mitigate this problem, with LLMs serving as advanced aids subject to ongoing real-world scrutiny.

Equitable access to these tools also requires attention. Many smaller hospitals and safety-net facilities face resource constraints that hinder deployment of advanced AI software.^64^ Models capable of running locally, securely, and at reasonable cost could help close this gap. Data privacy, liability concerns, and regulatory oversight must keep pace with rapid innovations.^65^

Our review mapped some key challenges in the literature. Current evidence is narrow and fragile: most studies are retrospective, single-center, and occasionally exposed to data the models may have seen during pre-training. Proprietary APIs and undisclosed prompts block replication and external scrutiny. Metrics are reported on heterogeneous tasks, hampering comparison and masking over-fitting. Hallucinations and silent omissions remain an unresolved safety risk, especially where time-critical decisions are involved. Finally, future works should audit performance across sex, language, or socioeconomic features.

Limitations of this review. Our search was confined to PubMed and Google Scholar and limited to English-language papers available up to 30 January 2025; relevant work in other languages may have been missed. As a narrative review, we did not apply formal risk-of-bias tools or meta-analytic methods, and performance numbers were taken as reported. Rapid advances in both model capability and regulation mean that findings may date quickly.

Transformative shifts in stroke care typically arise from technologies that integrate into existing workflows and reduce morbidity and mortality.^66^ LLMs, if refined through transparent validation, may offer a new complement to acute triage and procedure documentation.^67,68^ AI-based language models may change how we interpret clinical data and guide decisions.

In conclusion, LLMs are promising research instruments for mining stroke documentation, but the jump to bedside use demands multisite validation, open benchmarks, bias audits, and robust human oversight. Until those conditions are met, these models should augment—not replace— expert clinical judgment in time-sensitive stroke care.

## Author Contributions

**Shelly Soffer**: Conceptualization, Methodology, Validation, Formal analysis, Investigation, Resources, Data Curation, Writing - Original Draft, Writing - Review & Editing, Visualization, Supervision, Project administration.

**Eyal Klang**: Conceptualization, Methodology, Software, Validation, Formal analysis, Investigation, Resources, Data Curation, Writing - Review & Editing, Supervision, Project administration.

**Aya Mudrik, Orly Efros, Mahmud Omar, Girish N Nadkarni:** Writing - Review & Editing, Validation, Supervision, Project administration.

## Competing interests

The authors declare no competing interests.

## Data availability statement

The datasets generated during and/or analyzed during the current study are available from the corresponding author on reasonable request.

## Funding sources

This research did not receive any specific grant from funding agencies in the public, commercial, or not-for-profit sectors.

## Notes

### Competing Interest Statement

The authors have declared no competing interest.

### Funding Statement

This study did not receive any funding

