## Supplementary Material for "Large Language Models in Stroke Management: A Review of the Literature"

**Supplement** **Materials**

**Search terms for PubMed and Google Scholar**

| **PubMed** | "stroke"[MeSH Terms] OR "stroke"[All Fields] OR "CVA"[All Fields] OR "stroke*"[All Fields] OR ("stroke"[MeSH Terms] OR "stroke"[All Fields] OR ("cerebrovascular"[All Fields] AND "accident"[All Fields]) OR "cerebrovascular accident"[All Fields]) OR ("apoplexies"[All Fields] OR "stroke"[MeSH Terms] OR "stroke"[All Fields] OR "apoplexy"[All Fields]) OR "poststroke"[All Fields] OR ("stroke"[MeSH Terms] OR "stroke"[All Fields] OR ("brain"[All Fields] AND "attack"[All Fields]) OR "brain attack"[All Fields])  AND  ("ChatGPT"[All Fields]) OR ("large language models"[All Fields]) OR ("OpenAI"[All Fields]) OR ("Microsoft bing"[All Fields]) OR ("google bard"[All Fields]) OR ("google gemini"[All Fields]) |
| --- | --- |
| **Google Scholar** | (stroke OR CVA OR "cerebrovascular accident")  AND  ("ChatGPT" OR "large language model" OR "large language models" OR OpenAI OR "Microsoft Bing" OR "Google Bard" OR "Google Gemini") |

**Supplementary Table 1:** Common limitations and future directions for LLM applications in stroke detection and imaging

| **Paper Reference** | **Limitations** | **Future Directions / Recommendations** |
| --- | --- | --- |
| **Lee** | - Published case reports may be atypical or incomplete  - Some mislocalizations (especially cerebellar or multi-lesion)  - No standard detail across all reports | - Future fine-tuning of LLMs specifically for neurology tasks  - Incorporating more consistent, structured patient-level data |
| **Kuzan** | - Retrospective design  - Limited specificity in precise lesion localization  - Only GPT-4V tested | - Prospective validation  - Technical improvements in image handling  - Further exploration of integration into clinical workflows |
| **Koyun** | - ChatGPT-4o had near-random specificity for AIS detection  - Only Claude 3.5 Sonnet and ChatGPT-4o tested  - Retrospective, relatively small sample | - More comprehensive model refinement  - Larger datasets to improve specificity  - Compare additional vision-enabled LLMs |
| **Wang X** | - Single-center, retrospective design  - Limited sample size  - Only GPT-3.5 and GPT-4 tested (no specialized medical LLM) | - Larger-scale or multicenter studies  - Further validation and refinement of prehospital screening approaches |

**Supplementary Table 2:** Common limitations and future directions for LLM applications in stroke data extraction

| **Paper Reference** | **Limitations** | **Future Directions / Recommendations** |
| --- | --- | --- |
| **Meddeb** | - Retrospective, multi-institutional but limited to two centers  - Language specificity (German or other) not explicitly stated, but likely  - Potential domain dependence (mechanical thrombectomy) | - Expand dataset and hospital sites  - Explore broader language support  - Develop more advanced prompts or structured data handling |
| **Goh** | - Small pilot (16 cases)  - Single-center, retrospective  - Limited variety in discharge summary styles | - Larger or multi-site validation  - Further refinement of local LLM prompts  - Integration into ongoing registry or audit systems |
| **Kim (PhenoFlow)** | - Design study with small set of case studies (only 4 neurologists for final testing)  - No broader user testing for generalizability | - Expand system to more sites or larger user groups  - Potential integration of more advanced statistical or AI-based interpretation |
| **Lehnen** | - Single expert as reference standard  - Only 30 external reports for validation  - Prompt optimization needed for certain tricky fields | - Larger validation cohorts  - Multicenter or prospective testing  - More refined prompt strategies |
| **Wang M** | - Single-center retrospective design  - Class imbalance (few hemorrhages)  - No fine-tuning, only default model parameters | - Possible multilingual or multicenter expansions  - Additional domain knowledge prompts or specialized training to handle complex procedural details |

**Supplementary Table 3:** Common limitations and future directions for LLM applications in stroke outcome prediction

| **Paper Reference** | **Limitations** | **Future Directions / Recommendations** |
| --- | --- | --- |
| **Phillips** | - Inconsistent imaging frequency  - Missing or irregular EHR data  - Smaller prospective sample  - Requires periodic recalibration if deployed in new settings | - Prospective trials of real-time risk alerts  - Protocols to confirm clinical impact on decision-making  - Adjust model for different hospital workflows |
| **Pedro T** | - Single-center, retrospective  - Exclusion of patients who died  - No direct imaging input for ChatGPT  - Potential biases in who was included | - Further large-scale or multicenter validation  - Potential to incorporate imaging data into future LLM-based approaches |

**Supplementary Table 4:** Common limitations and future directions for LLM applications in stroke rehabilitation

| **Paper Reference** | **Limitations** | **Future Directions / Recommendations** |
| --- | --- | --- |
| **Zhang** | - Single-case design  - Possible overgeneralization from one patient scenario  - Minor errors in ICF coding | - Domain-specific fine-tuning or training  - Evaluation on multiple real-world clinical cases for rehab prescriptions |
| **Cong** | - Retrospective, relying on AphasiaBank transcripts  - No direct mention of real-time or bedside feasibility | - Potential use with real-time speech input  - Explore domain-specific LLM fine-tuning for aphasia or neuro rehab |

**Supplementary Table 5:** Common limitations and future directions for LLM applications in stroke patient education

| **Paper Reference** | **Limitations** | **Future Directions / Recommendations** |
| --- | --- | --- |
| **Lam** | - Single example chat  - No quantitative performance metrics  - Potentially unusable for patients with severe aphasia or other communication barriers | - Further testing with larger user samples  - Strategies to accommodate deficits (e.g., voice-based or caregiver-based input)  - Collaboration with clinical triage systems |
| **Vora** | - Letter format, no formal evaluation  - Omitted key details (e.g., hypertension, strict treatment windows)  - Potential patient over-reliance on AI | - Inclusion of evidence-based guidelines and references  - Emphasize physician oversight when using ChatGPT for health information |
